# Effect of Opaganib on Supplemental Oxygen and Mortality in Patients with Severe SARS-CoV-2 Pneumonia

**DOI:** 10.1101/2022.06.12.22276088

**Authors:** Fernando Carvalho Neuenschwander, Ofra Barnett-Griness, Stefania Piconi, Yasmin Maor, Eduardo Sprinz, Nimer Assy, Oleg Khmelnitskiy, Nikita Lomakin, Boris Mikhailovich Goloshchekin, Ewelina Nahorecka, Adilson Joaquim Westheimer Calvacante, Anastasia Ivanova, Sergey Vladimirovich Zhuravel, Galina Yurevna Trufanova, Stefano Bonora, Amer Saffoury, Ami Mayo, Yury Shvarts, Giuliano Rizzardini, Rogerio Sobroza de Mello, Janaina Pilau, Alexey Klinov, Benjamin Valente-Acosta, Oleg Olegovich Burlaka, Natalia Bakhtina, Maskit Bar-Meir, Ivan Nikolaevich Shishimorov, Jose Oñate-Gutierrez, Cristian Ivan Garcia Rincon, Tatiana Ivanovna Martynenko, Ludhmila Abrahão Hajjar, Ana Carolina Nazare de Mendonca Procopio, Krzystof Simon, Walter Gabriel Chaves Santiago, Adam Fronczak, Conrado Roberto Hoffmann Filho, Osama Hussein, Vladimir Aleksandrovich Martynov, Guido Chichino, Piotr Blewaska, Jacek Wroblewski, Sergio Saul Irizar Santana, Andres Felipe Ocampo Agudelo, Adam Barczyk, Rachael L. Gerlach, Eppie Campbell, Aida Bibliowicz, Reza Fathi, Patricia Anderson, Gilead Raday, Michal Klein, Clara Fehrmann, Gina Eagle, Vered Katz Ben-Yair, Mark L. Levitt

**Author notes:** Corresponding Author: Mark L. Levitt, RedHill Biopharma Ltd., 21 Ha’arba’a St., Tel Aviv, Israel 647392. This article has an online data supplement, which is accessible from this issue’s table of content online at www.atsjournals.org. **Contributions:** Conceptualization - Ofra Barnett-Griness, Gilead Raday, Gina Eagle, Vered Katz Ben-Yair, Mark L. Levitt Methodology - Ofra Barnett-Griness, Michal Klein, Gilead Raday, Gina Eagle, Vered Katz Ben-Yair, Mark L. Levitt Data curation - Ofra Barnett-Griness, Aida Bibliowicz, Eppie Campbell, Rachael L. Gerlach, Gilead Raday, Gina Eagle, Clara Fehrmann, Vered Katz Ben-Yair, Mark L. Levitt, Reza Fathi, Patricia Anderson Formal analysis - Ofra Barnett-Griness, Michal Klein, Gina Eagle, Vered Katz Ben-Yair, Mark L. Levitt, Gilead Raday Writing - original draft - Gina Eagle, Vered Katz Ben-Yair, Mark L. Levitt Writing - reviewing and editing - Ofra Barnett-Griness, Aida Bibliowicz, Reza Fathi, Patricia Anderson, Gilead Raday, Gina Eagle, Vered Katz Ben-Yair, Mark L. Levitt, Michal Klein, Clara Fehrmann, Rachael L. Gerlach, Eppie Campbell, Neuenschwander, Piconi, Maor, Sprinz, Assy, Khmelnitskiy, Lomakin, Goloshchekin, Nahorecka, Calvacante, Ivanova, Zhuravel, Trufanova, Bonora, Saffoury, Mayo, Shvarts, Rizzardini, Sobroza de Mello, Pilau, Klinov, Valente-Acosta, Burlaka, Bakhtina, Bar-Meir, Shishimorov, Oñate-Gutierrez, Rincon, Martynenko, Abrahão Hajjar, Procopio, Santiago, Simon, Filho, Fronczak, Hussein, Martynov, Agudelo, Blewaska, Chichino, Santana, Wroblewski, Barczyk Principal Investigator - Mark L. Levitt Investigators - Neuenschwander, Piconi, Maor, Sprinz, Assy, Khmelnitskiy, Lomakin, Goloshchekin, Nahorecka, Calvacante, Ivanova, Zhuravel, Trufanova, Bonora, Saffoury, Mayo, Shvarts, Rizzardini, Sobroza de Mello, Pilau, Klinov, Valente-Acosta, Burlaka, Bakhtina, Bar-Meir, Shishimorov, Oñate-Gutierrez, Rincon, Martynenko, Abrahão Hajjar, Procopio, Santiago, Simon, Filho, Fronczak, Hussein, Martynov, Agudelo, Blewaska, Chichino, Santana, Wroblewski, Barczyk. **Additional Contributions:** The authors would like to acknowledge the patients who participated in the study, as well as the dedicated healthcare professionals who cared for them at risk of their own lives. The authors would like to thank Dr. W. Todd Penberthy for his assistance in manuscript writing and editing. **Funding:** RedHill Biopharma, Ltd.

## Abstract

**Rationale:** There are few treatment options for severe COVID-19 pneumonia. Opaganib is an oral treatment under investigation.

**Objective:** Evaluate opaganib treatment in hospitalized patients with severe COVID-19 pneumonia.

**Methods:** A randomized, placebo-controlled, double-blind phase 2/3 trial was conducted in 60 sites worldwide from August 2020 to July 2021.

Patients received either opaganib (n=230; 500mg twice daily) or matching placebo (n=233) for 14 days.

**Main Outcome Measurements:** Primary outcome was the proportion of patients no longer requiring supplemental oxygen by day 14. Secondary outcomes included changes in the World Health Organization Ordinal Scale for Clinical Improvement, viral clearance, intubation, and mortality at 28- and 42-days.

**Main Results:** Pre-specified primary and secondary outcome analyses did not demonstrate statistically significant benefit (except for time to viral clearance). Post-hoc analysis revealed the fraction of inspired oxygen (FiO_2_) at baseline was prognostic for opaganib treatment responsiveness and corresponded to disease severity markers. Patients with FiO_2_ levels at or below the median value (≤60%) had better outcomes after opaganib treatment (n=117) compared to placebo (n=134). The proportion of patients with ≤60% FIO2 at baseline that no longer required supplemental oxygen (≥24 hours) by day 14 of opaganib treatment increased (76.9% vs 63.4%: p-value =0.033). There was a 62.6% reduction in intubation/mechanical ventilation (6.84% vs 17.91%; p-value=0.012) and a clinically meaningful 62% reduction in mortality (5.98% vs 16.7%; p-value=0.019) by day 42. No new safety concerns observed.

**Conclusions:** Post-hoc analysis supports opaganib benefit in COVID-19 severe pneumonia patients that require lower supplemental oxygen (≤60% FiO2). Further studies are warranted.

**Trial registration number:** NCT04467840

## INTRODUCTION

Severe Acute Respiratory Syndrome Coronavirus 2 (SARS-CoV-2) has caused more than five million deaths by coronavirus disease-19 (COVID-19) worldwide.^1^ Patients infected with SARS-CoV-2 may progress to acute respiratory distress syndrome followed by death. The global need for therapeutics to safely treat severe COVID-19 has becomes more urgent with recent increases in breakthrough SARS-CoV-2 infections due to emerging viral variants and continued vaccine hesitancy.^2^

Currently, Intravenous remdesivir is the only therapeutic approved for use in cases of severe COVID-19, by the European Medicines Agency (EMA) and the Food and Drug Administration (FDA).^3^ In addition, other more recent studies have not found remdesivir to be effective.^4^ Dexamethasone has been granted approval in the United Kingdom for oral or IV treatment for COVID-19 while molnupiravir and Paxlovid were recently approved by the FDA for outpatients within 5 days of symptom onset with some safety concerns.^5–10^

Opaganib ([3-(4-chlorophenyl)-adamantane-1-carboxylic acid (pyridin-4-ylmethyl)amide, hydrochloride salt]) is an oral, first in class sphingosine kinase 2 (SK2) selective inhibitor.^11^ SK2 is a molecular target due to its critical role in sphingolipid metabolism, Including the replication-transcription complex (RTC) of certain RNA viruses.^12^

Preclinical studies of the Chikungunya virus, a coding strand single strand RNA virus (like SARS-CoV-2), determined that inhibition of SK2 with opaganib markedly inhibited viral transcription.^12^ In addition, our pre-clinical studies have demonstrated that opaganib is a potent inhibitor of SARS-CoV-2 replication.

Importantly, SK2 is likely unaffected by viral mutations, including those of the viral spike protein. For example, omicron has over 30 mutations in the spike protein, diminishing the effectiveness of current vaccines that target these epitopes.^13–15^ Omicron and future variants have the potential to evade vaccine induced immunity.

In a *Pseudomonas aeruginosa* pneumonia model, opaganib decreased lung injury and associated inflammation.^16^ Thus, inhibition of SK2 by opaganib may provide therapeutic benefit in reducing lung inflammatory injury.

Given the potential anti-viral and anti-inflammatory properties of opaganib, we conducted 2 clinical trials to evaluate opaganib for the treatment of SARS-CoV-2 infection in hospitalized patients with COVID-19 pneumonia. The clinical trials included a completed phase 2a proof of concept study, and a completed phase 2/3 global study with results of the latter summarized in this report.^17–19^

The phase 2a study (n=42, non-powered) evaluated supplemental oxygen requirements, time to discharge, and safety in hospitalized patients with COVID-19 pneumonia requiring supplemental oxygen. Patients receiving oral opaganib showed a numerical trend for less supplemental oxygen requirement on treatment, resulting in earlier hospital discharge, with no notable safety issues.^19^ These results led us to evaluate a 14-day course of opaganib therapy for reduction in the need for supplemental oxygen and improvement in the clinical status of hospitalized patients with COVID-19 pneumonia in the phase 2/3 study, presented herein.

## METHODOLOGY

### STUDY SETTINGS AND TRIAL DESIGN

This clinical trial was a multicenter, phase 2/3, randomized, double-blind, placebo-controlled, opaganib treatment study involving patients diagnosed with COVID-19 infection defined by eligibility criteria generally aligned with the World Health Organization (WHO) Ordinal Scale for Clinical Improvement level 5. Recruitment was from August 2020 to July 2021 and the study was performed at 57 locations in 7 countries (NCT04467840). Specifically, eligible patients had pneumonia secondary to SARS-CoV-2 based on radiographic evidence via chest X-ray or CT scan, and patients required supplemental oxygen by high flow nasal cannula, positive pressure ventilation or non-rebreather face masks. For a short period, patients with simple face masks and an oxygen flow rate greater than 5L/min were allowed, due to lack of adequate high flow devices at several sites. Patients were required to be hospitalized at baseline day 1. The trial complied with the Declaration of Helsinki, the International Conference on Harmonization Guidelines for Good Clinical Practice, and applicable local regulations. The protocol was approved by the ethics committees at all participating centers. All patients provided written informed consent before study entry.

### RANDOMIZATION AND INTERVENTION

Patients received local SoC) and were randomized to receive either 2 x 250 mg opaganib capsules (500 mg) every 12 hours or matching placebo, in a randomization ratio of 1:1. Stratification factors were: whether the patients met three or more high risk parameters for COVID-19 outcomes at baseline (yes or no) and whether SoC treatment has established efficacy (yes or no). The risk parameters list as well as the list of SoC treatments considered efficacious are provided in supplemental material 1 and 2 respectively. Treatment assignments were blinded to the patient, investigator, and hospital staff, as well as the sponsor. Study drug was administered for 14 days, participants were followed for 42 days from their first dose.

### PRIMARY AND SECONDARY OUTCOMES

The primary outcome was a measurement of the proportion of patients no longer requiring supplemental oxygen by the end of treatment day 14, which was defined per patient in a binary manner as “success” or “failure”. If a patient no longer required supplemental oxygen, for at least 24 hours by treatment day 14, then this was considered a success, provided that resumption of supplemental oxygen did not occur through day 42. Death or withdrawal from the study was considered as failure regardless of previous success.

A total of nine efficacy secondary outcomes were evaluated, including viral clearance, intubation and mortality. For further information on methodology, please see the methodology section in the online data supplement.

Adverse events (AEs) and serious AEs (SAEs) were evaluated with respect to vital signs, laboratory parameters (chemistry and hematology), electrocardiograms, and incidence rates of treatment-emergent AEs (TEAEs) and SAEs.

## RESULTS

### PATIENT DEMOGRAPHICS AND CLINICAL CHARACTERISTICS

A total of 588 patients were initially screened, and 475 eligible patients were randomized (237 to opaganib, 238 to placebo; see consort diagram, Figure 1). There were 113 screen failures, the reasons for screen failure are presented in Table E1 in the online data supplement. Study treatment was completed by 80.9% in the opaganib arm (n=186) and 79.0% in the placebo arm (n=186). Investigator decision to stop study drug was made for 0.6% (3 out of 475) patients, one in the opaganib arm and 2 in the placebo arm. A total of 36 (15.6%) patients in the opaganib arm and 40 (17.4%) patients in the placebo arm discontinued study due to death. Two deaths, both in the placebo arm, occurred after completion of the 42 day follow up.

**Figure 1.**
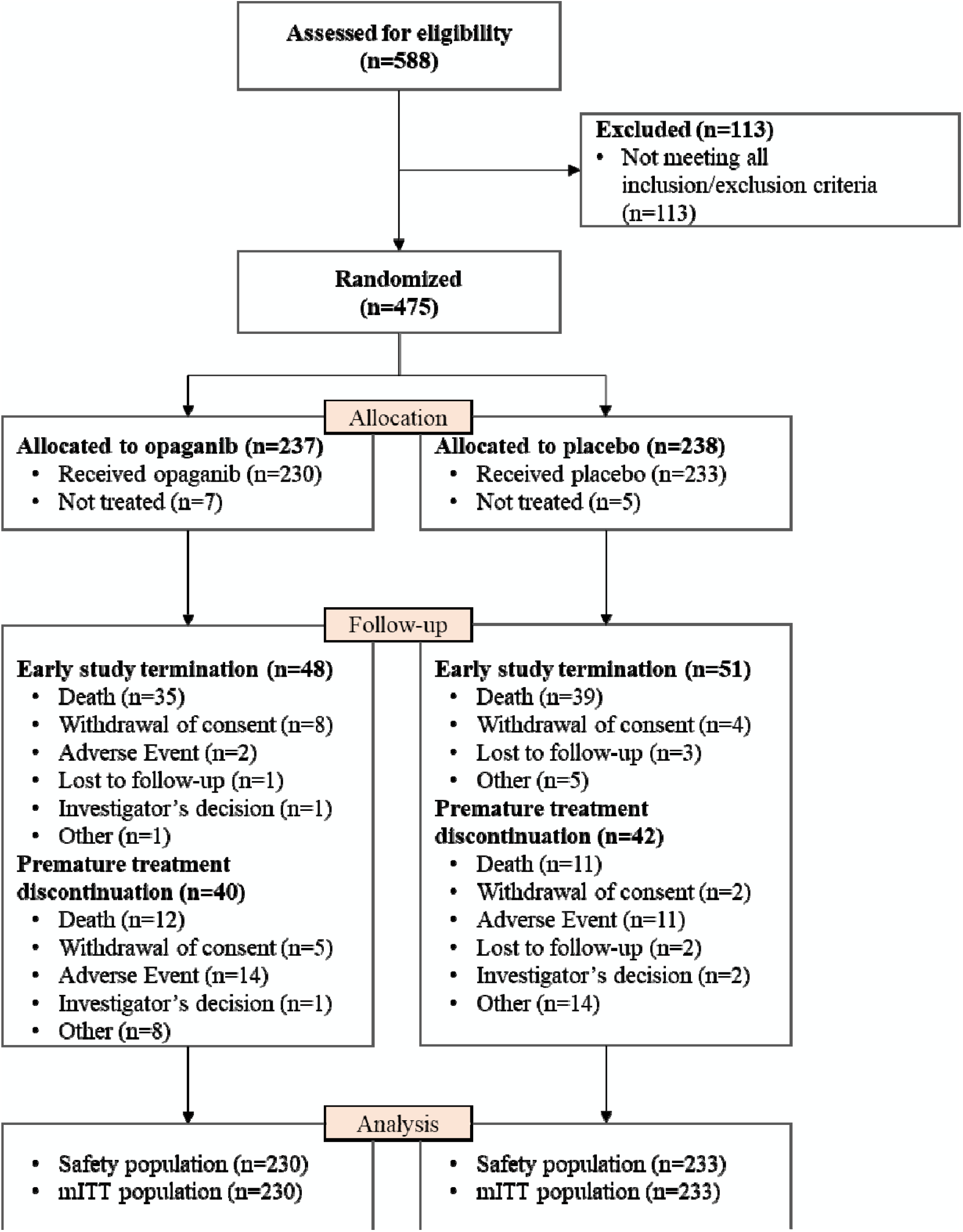
Consort flow diagram. mITT=modified Intention to treat, all subjects who were randomized and treated with at least one dose (even partial) of study drug.

Patient demographics and characteristics were similar between treatment arms as shown in Table 1. There were no meaningful differences in age, smoking status, or HbA1C status. The type of oxygen delivery device (i.e., non-invasive positive pressure [BiPAP or CPAP], high flow nasal cannula [HFNC], or face mask [with or without reservoir]), was similar between treatment groups. Other characteristics at baseline were similar between treatment arms are shown. In addition, weight, height, and body mass index (BMI) were also similar between treatment arms as were concomitant medications. Effective SoC medications for COVID-19 treatments included glucocorticoids (predominantly dexamethasone), remdesivir, and COVID-19 convalescent plasma. Glucocorticoids were the most used, with 94% of all study patients receiving glucocorticoids.

**Table 1.**
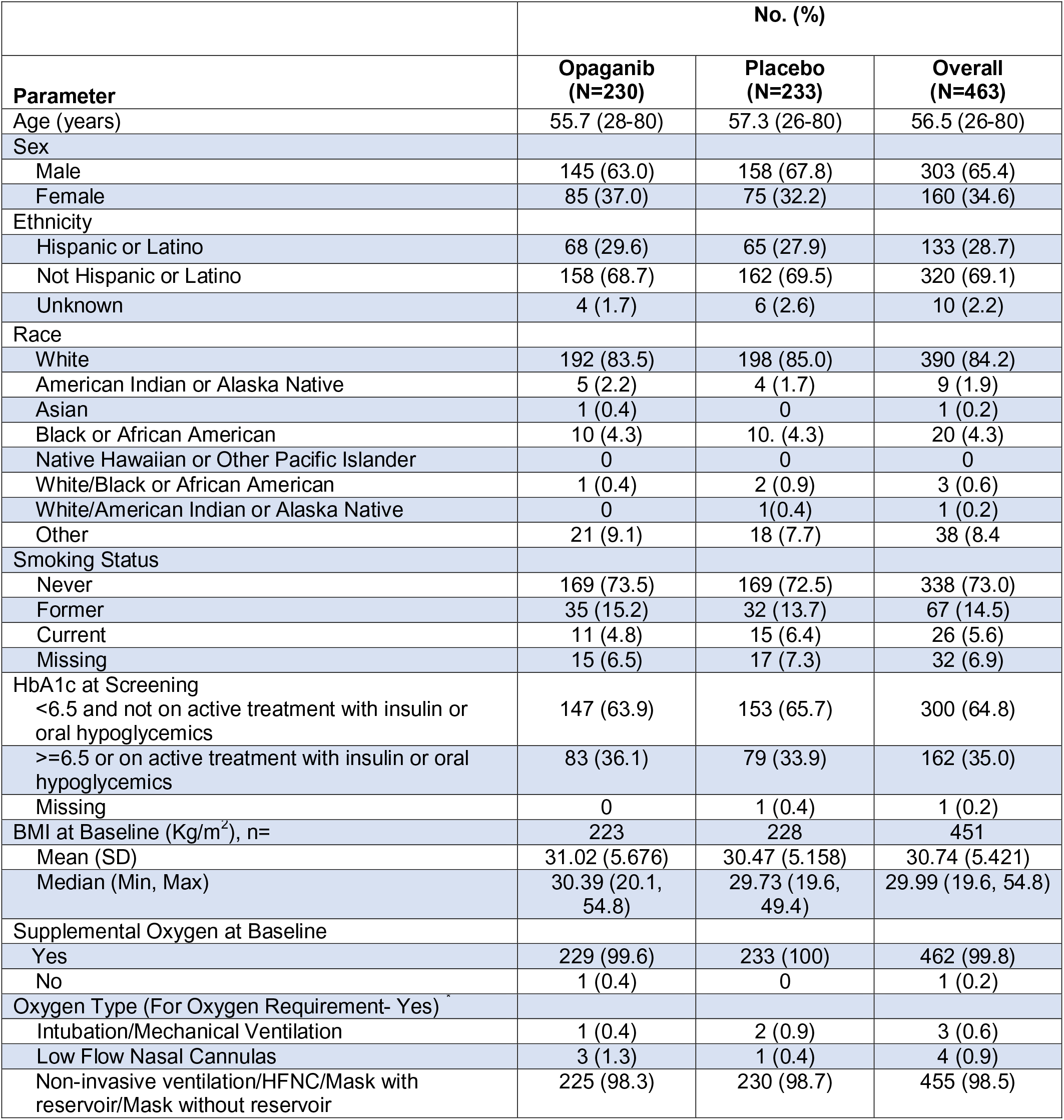
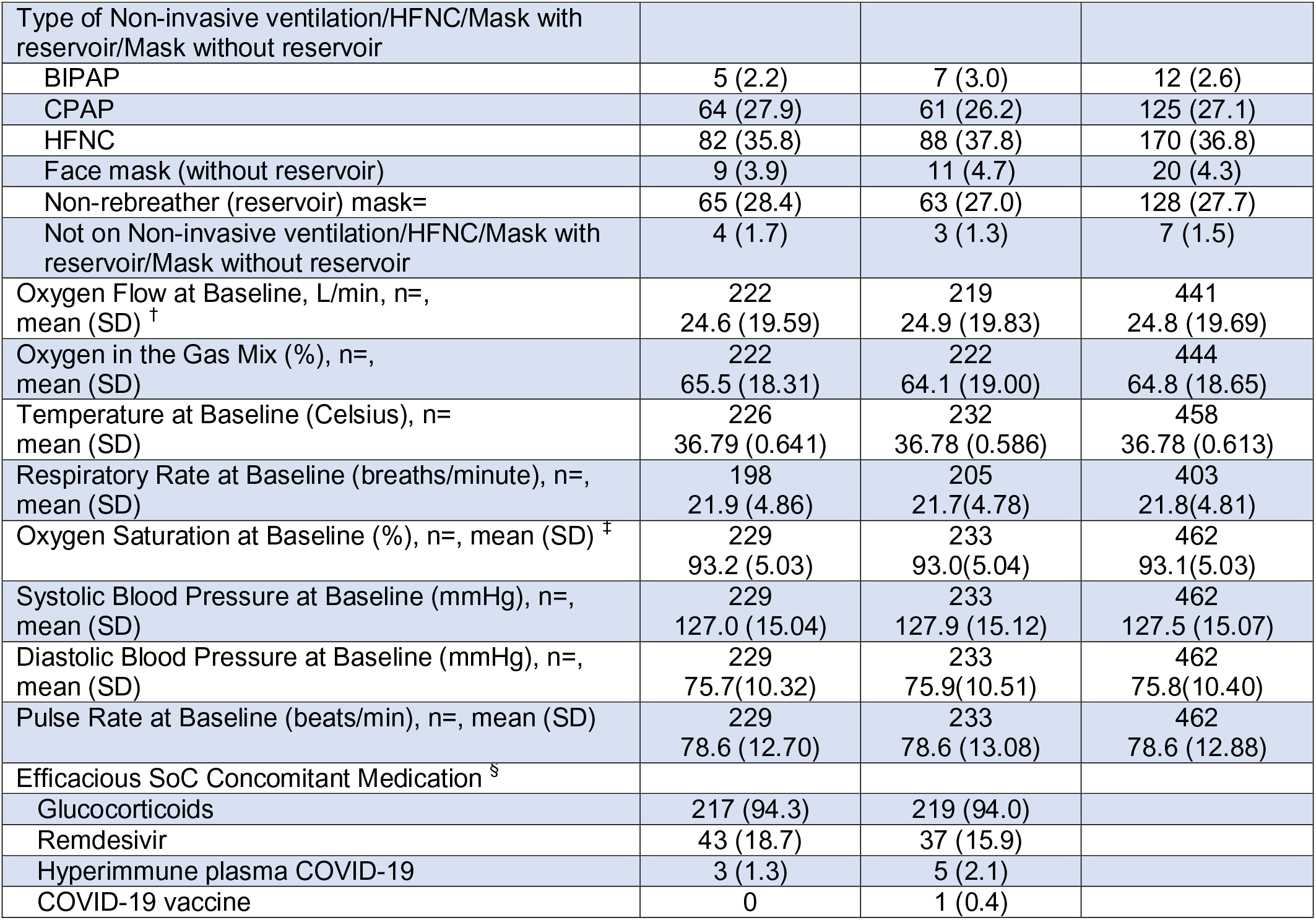
Patient demographics and characteristics. All baseline oxygen variables are taken from the last observation prior to exposure or if time not available from exposure date. *Percentages are calculated from patients who required oxygen at baseline. ^†^Baseline oxygen flow was only collected among patients not intubated at baseline who required oxygen. ^‡^Patients who could not be taken off of oxygen to measure their saturation on room air, had measurements taken while on supplemental oxygen. ^§^Concomitant medications received at any timepoint during the treatment period of 14 days

|  | No. (%) |  |  |
| --- | --- | --- | --- |
| Parameter | Opaganib<br>(N=230) | Placebo<br>(N=233) | Overall<br>(N=463) |
| Age (years) | 55.7 (28-80) | 57.3 (26-80) | 56.5 (26-80) |
| Sex |  |  |  |
| Male | 145 (63.0) | 158 (67.8) | 303 (65.4) |
| Female | 85 (37.0) | 75 (32.2) | 160 (34.6) |
| Ethnicity |  |  |  |
| Hispanic or Latino | 68 (29.6) | 65 (27.9) | 133 (28.7) |
| Not Hispanic or Latino | 158 (68.7) | 162 (69.5) | 320 (69.1) |
| Unknown | 4 (1.7) | 6 (2.6) | 10 (2.2) |
| Race |  |  |  |
| White | 192 (83.5) | 198 (85.0) | 390 (84.2) |
| American Indian or Alaska Native | 5 (2.2) | 4 (1.7) | 9 (1.9) |
| Asian | 1 (0.4) | 0 | 1 (0.2) |
| Black or African American | 10 (4.3) | 10 (4.3) | 20 (4.3) |
| Native Hawaiian or Other Pacific Islander | 0 | 0 | 0 |
| White/Black or African American | 1 (0.4) | 2 (0.9) | 3 (0.6) |
| White/American Indian or Alaska Native | 0 | 1 (0.4) | 1 (0.2) |
| Other | 21 (9.1) | 18 (7.7) | 38 (8.4) |
| Smoking Status |  |  |  |
| Never | 169 (73.5) | 169 (72.5) | 338 (73.0) |
| Former | 35 (15.2) | 32 (13.7) | 67 (14.5) |
| Current | 11 (4.8) | 15 (6.4) | 26 (5.6) |
| Missing | 15 (6.5) | 17 (7.3) | 32 (6.9) |
| HbA1c at Screening |  |  |  |
| <6.5 and not on active treatment with insulin or oral hypoglycemics | 147 (63.9) | 153 (65.7) | 300 (64.8) |
| >=6.5 or on active treatment with insulin or oral hypoglycemics | 83 (36.1) | 79 (33.9) | 162 (35.0) |
| Missing | 0 | 1 (0.4) | 1 (0.2) |
| BMI at Baseline (Kg/m<sup>2</sup>), n= | 223 | 228 | 451 |
| Mean (SD) | 31.02 (5.676) | 30.47 (5.158) | 30.74 (5.421) |
| Median (Min, Max) | 30.39 (20.1, 54.8) | 29.73 (19.6, 49.4) | 29.99 (19.6, 54.8) |
| Supplemental Oxygen at Baseline |  |  |  |
| Yes | 229 (99.6) | 233 (100) | 462 (99.8) |
| No | 1 (0.4) | 0 | 1 (0.2) |
| Oxygen Type (For Oxygen Requirement- Yes) |  |  |  |
| Intubation/Mechanical Ventilation | 1 (0.4) | 2 (0.9) | 3 (0.6) |
| Low Flow Nasal Cannulas | 3 (1.3) | 1 (0.4) | 4 (0.9) |
| Non-invasive ventilation/HFNC/Mask with reservoir/Mask without reservoir | 225 (98.3) | 230 (98.7) | 455 (98.5) |
| Type of Non-invasive ventilation/HFNC/Mask with reservoir/Mask without reservoir |  |  |  |
| BIPAP | 5 (2.2) | 7 (3.0) | 12 (2.6) |
| CPAP | 64 (27.9) | 61 (26.2) | 125 (27.1) |
| HFNC | 82 (35.8) | 88 (37.8) | 170 (36.8) |
| Face mask (without reservoir) | 9 (3.9) | 11 (4.7) | 20 (4.3) |
| Non-rebreather (reservoir) mask= | 65 (28.4) | 63 (27.0) | 128 (27.7) |
| Not on Non-invasive ventilation/HFNC/Mask with reservoir/Mask without reservoir | 4 (1.7) | 3 (1.3) | 7 (1.5) |
| Oxygen Flow at Baseline, L/min, n=, mean (SD) <sup>†</sup> | 222<br>24.6 (19.59) | 219<br>24.9 (19.83) | 441<br>24.8 (19.69) |
| Oxygen in the Gas Mix (%), n=, mean (SD) | 222<br>65.5 (18.31) | 222<br>64.1 (19.00) | 444<br>64.8 (18.65) |
| Temperature at Baseline (Celsius), n=, mean (SD) | 226<br>36.79 (0.641) | 232<br>36.78 (0.586) | 458<br>36.78 (0.613) |
| Respiratory Rate at Baseline (breaths/minute), n=, mean (SD) | 198<br>21.9 (4.86) | 205<br>21.7(4.78) | 403<br>21.8(4.81) |
| Oxygen Saturation at Baseline (%), n=, mean (SD) <sup>‡</sup> | 229<br>93.2 (5.03) | 233<br>93.0(5.04) | 462<br>93.1(5.03) |
| Systolic Blood Pressure at Baseline (mmHg), n=, mean (SD) | 229<br>127.0 (15.04) | 233<br>127.9 (15.12) | 462<br>127.5 (15.07) |
| Diastolic Blood Pressure at Baseline (mmHg), n=, mean (SD) | 229<br>75.7(10.32) | 233<br>75.9(10.51) | 462<br>75.8(10.40) |
| Pulse Rate at Baseline (beats/min), n=, mean (SD) | 229<br>78.6 (12.70) | 233<br>78.6 (13.08) | 462<br>78.6 (12.88) |
| Efficacious SoC Concomitant Medication <sup>§</sup> |  |  |  |
| Glucocorticoids | 217 (94.3) | 219 (94.0) |  |
| Remdesivir | 43 (18.7) | 37 (15.9) |  |
| Hyperimmune plasma COVID-19 | 3 (1.3) | 5 (2.1) |  |
| COVID-19 vaccine | 0 | 1 (0.4) |  |

### PRIMARY AND SECONDARY EFFICACY OUTCOMES

Primary analysis of the mITT population revealed 139 (60.4%) of patients receiving opaganib were no longer receiving supplemental oxygen by day 14 as compared to 132 (56.7%) of patients receiving placebo (p-value 0.391). While opaganib was numerically superior to placebo, pre-specified analyses of primary and endpoint did not demonstrate a statistically significant treatment differences in benefit in the mITT population as shown in Table 2. A consistent albeit small numerical benefit was shown across the secondary endpoints, including measures of the change days until improvement in the WHO Ordinal Scale status (i.e., time to room air), the time (days) to low oxygen flow via nasal cannula, the time (days) to hospital discharge, the proportion of patients requiring intubation and ventilation by day 42, and the mortality percentage by day 42 and percent viral clearance by day 14 as shown Tables E2 and E3 in the online data supplement. Of note, the secondary endpoint requiring serial measurements of temperature could not be evaluated due to the volume of missing data points.

**Table 2.** Primary endpoint analysis, percentage of patients no longer requiring supplemental oxygen, for at least 24 hours by study day 14. *Success indicates that a patient was no longer receiving supplemental oxygen for at least 24 hours by day 14. ^†^p-value from Cochran Mantel-Haenzel test using the study stratification factors used for randomization, and corresponding stratified proportion difference with 95%CI. All reported p-values are two sided. Patients can only be on one failure category. ^‡^Patients who die within 42-days or are LTFU or in need of oxygen up to 42 days are regarded as Failure.

| Parameter | No. (%) |  | Opaganib vs Placebo |
| --- | --- | --- | --- |
|  | Opaganib (N=230) | Placebo (N=233) |  |
| Patients No Longer Receiving Supplemental Oxygen ("Success" <sup>*</sup>), n (%) | 139 (60.43) | 132 (56.65) |  |
| Difference <sup>†</sup> |  |  | +3.78 |
| 95% CI for the difference <sup>†</sup> |  |  | -5.19, 12.75 |
| p-value <sup>†</sup> |  |  | 0.391 |
| "Failure" <sup>‡</sup> | 91 (39.57) | 101 (43.35) |  |
| Due to need of supplemental oxygen at Day 14, n (%) | 70 (30.43) | 75 (32.19) |  |
| Due to death up to day 14, n (%) | 18 (7.83) | 21 (9.01) |  |
| Due to Lost to follow up by Day 14, n (%) | 3 (1.30) | 4 (1.72) |  |
| Due to missing status at Day 42 with prior success, n (%) | 0 | 1 (0.43) |  |

Since opaganib previously demonstrated potent anti-viral efficacy *in vitro* in a SARS-CoV-2 model (data on file, RedHill Biopharma), one of the secondary outcomes evaluated was time to viral clearance. In a pre-specified analysis, opaganib demonstrated a nominally significant improvement in time to viral clearance as compared to placebo, with a hazard ratio of 1.34 and a p-value of 0.043 as shown in Figure 2 (and Table E3 in the online data supplement). The median time to viral clearance was 10 days for the opaganib treated arm compared to >14 days for the placebo arm.

**Figure 2.**
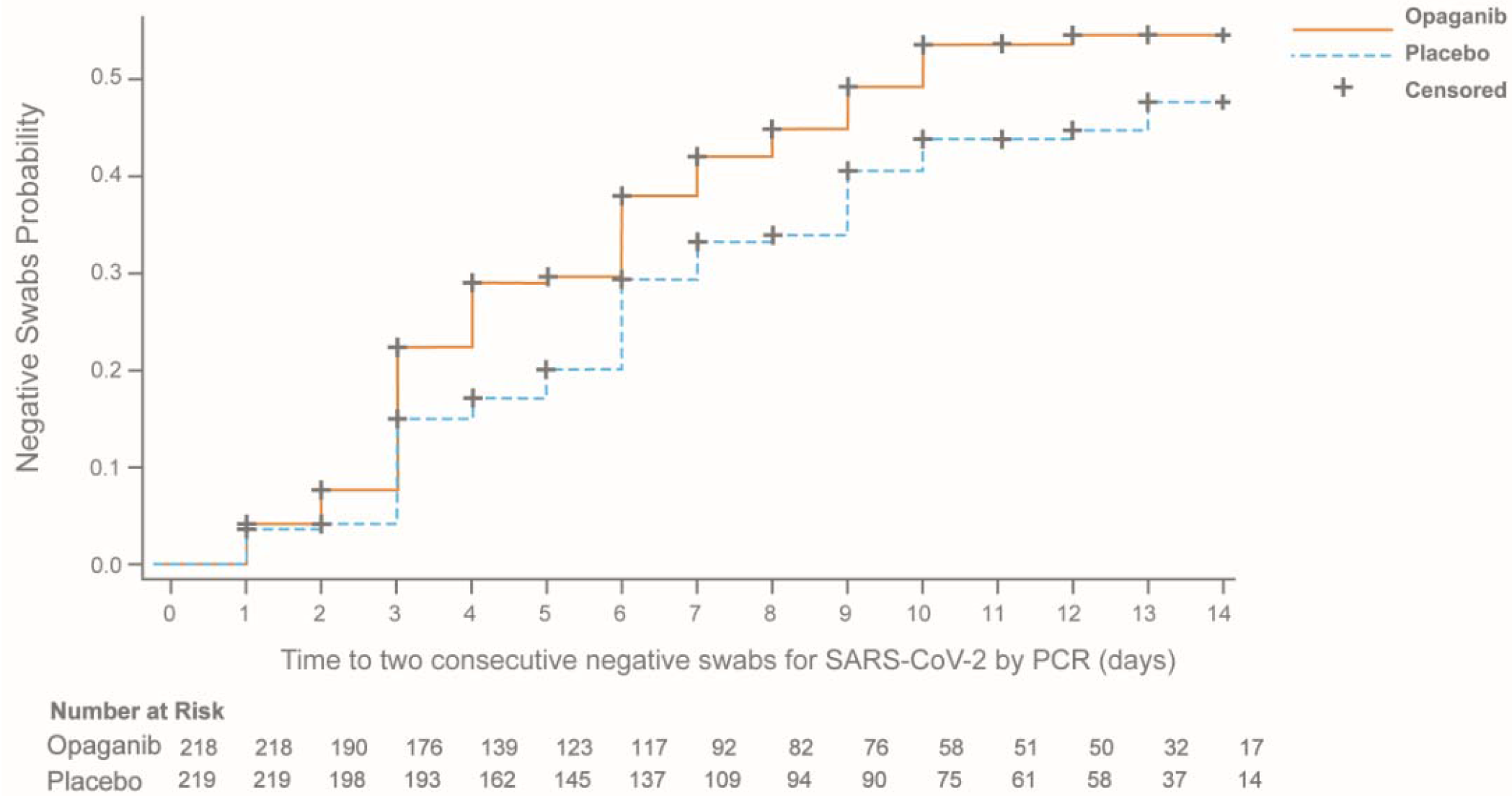
Kaplan-Meier curve of cumulative incidence for time to two consecutive negative swabs for SARS-CoV-2 by PCR, at least 24hrs. apart, in the mITT population that were PCR positive for SARS-CoV-2 at screening. Twenty-six (26) patients that were excluded from this analysis had an eligibility PCR up to 7 days prior to screening, but not at screening.

Pre-specified strata analysis revealed that opaganib treatment reduced mortality by day 28 (4.7% vs 21.3%; p-value=0.024) and day 4 compared to patients receiving remdesivir and corticosteroid SoC (n=90) and by day 42 (7.0% vs 23.4%; p-value=0.034; Table E4 in the online data supplement). Furthermore, time to recovery as defined by improvement to a score of 1 or less on the WHO Ordinal Scale at 14 days of treatment was assessed in the mITT (n=463) analysis set (Table E5 and Figure E1 in the online data supplement), as a pre-specified exploratory objective. Opaganib treatment reduced the time until recovery with 86 (37.4%) opaganib-treated patients vs 65 (27.9%) in the placebo arm recovering by day 14 (p-value = 0.013, HR 1.49).

### POST-HOC EFFICACY ANALYSIS

A pre-specified sensitivity analysis revealed that the sub-population comprised of patients treated by either high flow nasal cannula, non-mechanical positive pressure ventilation, or reservoir face masks at baseline demonstrated a reduced treatment benefit with opaganib as shown in Table E6 in the online data supplement. Thus, we decided to perform two post hoc analyses of opaganib versus placebo using the other oxygenation parameter that had been collected: one for patients at or below and one for patients above the median value of the FiO_2_ (60%).

Post-hoc analysis of the subpopulation requiring relatively lower FiO_2_ (≤60%; n=251; Table 3) at baseline demonstrated an appreciable positive opaganib treatment benefit with no benefit observed for patients requiring higher FiO_2_ (>60%; n=193) at baseline. Primary outcome analysis demonstrated a 21.3% relative increase in patients no longer requiring supplemental oxygen vs placebo (76.9% vs 63.4%, nominal p-value p=0.033). This post hoc primary endpoint analysis was supported by the outcomes for the secondary endpoints in this subpopulation, associated with clinical outcomes and supplemental oxygen requirements, with nominally significant p-values, including for (1) changes in the WHO Ordinal Scale (21% relative increase), (2) the need for intubation/mechanical ventilation (61.8% reduction), and (3) reduction in overall mortality by day 42 (61.8% reduction). The FiO2 correlated well with the SpO2:FiO2 ratio, a validated predictor for Acute Respiratory Distress Syndrome (R= −0.93).

**Table 3.** Post-hoc primary and secondary outcome analyses in the subpopulation requiring ≤60% FiO2 at baseline. N (and n) =number; CI = Confidence Interval. * Success indicating that a patient no longer received supplemental oxygen for at least 24 hours by Day 14. ^†^Nominal p-value from Cochran Mantel-Haenszel test using the study stratification factors used for randomization, and corresponding stratified proportion difference with 95%CI. All reported p-values are nominal, two sided. Patients can only be on one failure category. ^‡^Success was defined as subject who reached improvement of at least two points on the WHO Ordinal Scale by Day 14, maintained by end of study (EOS). ^§^Estimated using the Kaplan-Meier estimator. Standard error of the medinas were estimated using the bootstrap method with 10,000 sample size. ^ll^Patients who died within 42-days or were LTFU or in need of oxygen up to 42 days have been regarded as failure. For secondary outcomes failure was defined for any requirement of intubation and mechanical ventilation or death without intubation by day 42. Early termination of study (or failure to complete EOS visit) is also considered as failure. **Mortality (“failure”) is assessed by treatment day 42 (including). Same results were demonstrated for Day 28. Any early termination/ missing survival status at EOS visit is also regarded as failure for the primary analysis of this endpoint.

|  | No. (%) |  |  |
| --- | --- | --- | --- |
|  | Opaganib<br>(N=117) | Placebo<br>(N=134) | Opaganib vs<br>Placebo |
| **Primary Outcomes** |  |  |  |
| Patients No Longer Receiving Supplemental Oxygen ("Success") * | 90 (76.9) | 85 (63.4) |  |
| Difference <sup>†</sup> |  |  | 13.49 |
| 95% CI for the difference <sup>†</sup> |  |  | 2.32, 24.66 |
| P-value <sup>†</sup> |  |  | 0.033 |
| "Failure" | 27 (23.08) | 49 (36.57) |  |
| Due to need of supplemental oxygen at day 14 | 24 (20.51) | 32 (23.88) |  |
| Due to death up to day 14 | 3 (2.56) | 13 (9.70) |  |
| Due to Lost to follow up by Day 14 | 0 | 3 (2.24) |  |
| Due to missing status at Day 42 with prior success | 0 | 1 (0.75) |  |
| **Secondary Outcomes** |  |  |  |
| Patients with an Improvement of 2 or more on the WHO Ordinal Scale Compared to Baseline by day 14 <sup>‡</sup> | 93 (79.49) | 88 (65.67) |  |
| Difference |  |  | 13.82 |
| Percentage change (Opaganib%/Placebo%*100-1) |  |  | 21 |
| P-value |  |  | 0.023 |
| Time to a score of $\leq 3$ on the WHO Ordinal Scale | | | |
| Number of Events | 93 (79.5) | 88 (65.7) |  |
| Kaplan-Meier Median (days) | 8.00 | 10.00 |  |
| 95% CI | 7.00 - 9.00 | 9.00 - 12.00 |  |
| P-value |  |  | 0.010 |
| Time to low oxygen flow via nasal cannula (from high flow nasal cannula or CPAP/BiPAP) |  |  |  |
| Number of Events | 102 (87.2) | 106 (79.1) |  |
| Kaplan-Meier Median (days) | 4.00 | 5.00 |  |
| 95% CI | 3.00 - 5.00 | 4.00 - 6.00 |  |
| P-value |  |  | 0.028 |
| Time to discharge by day 42 |  |  |  |
| Number of Events (%) | 110 (94.0) | 109 (81.3) |  |
| Kaplan-Meier Median (days) | 10.00 | 14.00 |  |
| 95% CI | 9.00 - 13.00 | 11.00 - 14.00 |  |
| P-value <sup>§</sup> |  |  | 0.004 |
| Patients requiring intubation and mechanical ventilation by day 42 | 8 (6.84) | 24 (17.91) |  |
| Difference |  |  | -11.07 |
| Percentage change (Opaganib%/Placebo%*100-1) |  |  | -61.8 |
| 95% CI | 2.26, 11.41 | 11.42, 24.40 | -19.01, -3.13 |
| Intubation Without death | 2 (1.71) | 3 (2.24) |  |
| Intubation with Death | 4 (3.42) | 13 (9.70) |  |
| Death without intubation | 1 (0.85) | 3 (2.24) |  |
| Early termination/missing data (alive without intubation) | 1 (0.85) | 5 (3.73) |  |
| P-value |  |  | 0.012 |
| Mortality due to any cause at day 42 ("Failure")<sup>**</sup> | 7 (5.98) | 21 (15.67) |  |
| Difference |  |  | -9.69 |
| Percentage change (Opaganib%/Placebo%*100-1) |  |  | -61.8 |
| 95% CI | 1.69, 10.28 | 9.52, 21.83 | -17.20, -2.18 |
| P-value |  |  | 0.019 |

A larger proportion of patients in this lower FiO_2_ cohort died in the placebo arm as compared to the opaganib arm (15.67% vs. 5.98% with % difference of -9.69; 95% CI - 17.21, -2.18; p-value 0.019; Table 3; Figure 3). Moreover, a larger proportion of patients requiring intubation and ventilation were observed in the placebo arm versus the opaganib arm by day 4 (17.91% vs. 6.84% with % difference of -11.0 7; 95% CI -19.01, -3.13; p-value 0.012).

**Figure 3.**
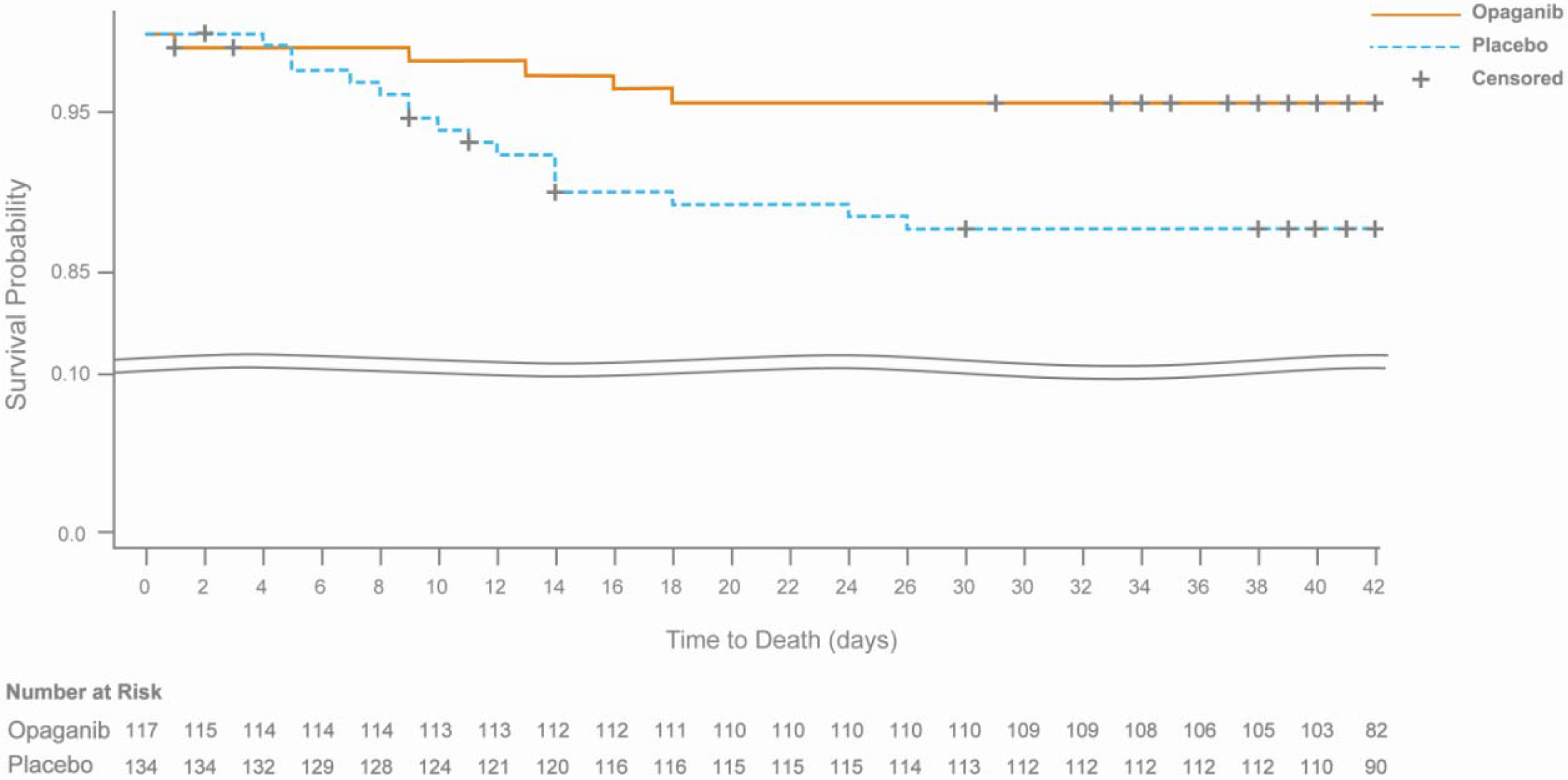
Kaplan-Meier curve of time to death through end of day 42 from mITT population with FiO2≤60% at baseline. Patients were censored at day 42 or at study termination if occurred before Day 42.

At baseline, lower median lymphocyte counts, and higher inflammatory median values were observed in the patients with ≥60% FiO_2_, thus supporting that FiO_2_ positively correlates with disease severity (Table 4). Consistent with the literature, higher LDH, lower lymphocyte count, higher D-Dimer, higher CRP and higher ferritin were significant risk factors for mortality. Advanced age and oxygen saturation at baseline were additional risk factors that positively correlated with worse outcomes, but higher FiO_2_ was ranked the second highest risk factor for mortality at day 42 in the mITT population (Table E7 in the online data supplement). No meaningful baseline differences in the values of these biomarkers were detectable when comparing the patients in the opaganib arm to the placebo arm in the low FiO_2_ group (Table E8 in the online data supplement).

**Table 4.** Biomarker medians by subpopulations (FiO2≤60% [low] and FiO2>60% [high] supplemental oxygen requirements at baseline). *Comparing FiO2≤60% vs. FiO2>60% subpopulations for imbalances.

| Marker | Low FiO<sub>2</sub> |  |  | High FiO<sub>2</sub> |  |  | p-value * |
| --- | --- | --- | --- | --- | --- | --- | --- |
|  | N | Median | Q1,Q3 | N | Median | Q1,Q3 |  |
| Lymphocytes (10<sup>9</sup>/L) | 249 | 0.990 | 0.71, 1.38 | 178 | 0.780 | 0.50, 1.20 | <.0001 |
| C Reactive Protein (mg/L) | 240 | 60.800 | 21.40, 153.40 | 186 | 102.800 | 38.67, 200.30 | 0.0005 |
| Ferritin (ug/L) | 228 | 666.95<br>0 | 370.17,<br>1297.50 | 167 | 1000.00<br>0 | 500.60,<br>2000.00 | 0.0008 |
| D-Dimer (ug/ml) | 240 | 0.450 | 0.18, 1.06 | 178 | 0.806 | 0.31, 1.79 | 0.0004 |
| Lactate Dehydrogenase (IU/L) | 230 | 361.15<br>0 | 290.60,<br>522.00 | 183 | 469.230 | 344.00,<br>644.00 | <.0001 |

The time from symptom onset to randomization in the mITT population was 11 days for both arms. Potential confounder effects on the mortality results derived from the baseline FiO_2_ subpopulation requiring ≤60% supplement oxygen was evaluated via Cox regression analysis (Table E9 of the online data supplement). We standardized (adjusted) each 42-day survival curve treatment group according to the distribution of respective confounders to potentially identify any effectors. Adjusted mortality rates were in close agreement with crude unadjusted estimates, thus confirming that it is unlikely that confounding effects contributed to the observed effect. In summary, Cox regression analysis of mortality outcomes suggested that the observed effects of opaganib treatment were independent of potential baseline characteristics confounders in this ≤60% FiO_2_ subpopulation.

### SAFETY ANALYSIS

Of 463 patients in the safety population who received at least one dose of study drug, 67.4% and 63.1% of patients receiving opaganib and placebo, respectively, experienced at least one treatment emergent adverse event (TEAE). The majority of the TEAEs were mild to moderate in severity. Treatment emergent serious adverse events (TESAEs) were experienced by 52/230 (22.6%) patients in the opaganib arm versus 52/233 (22.3%) patients in the placebo arm (Table 5). TEAEs with an outcome of death occurred in 36/230 (15.7%) versus 40/233 (17.2%) in the opaganib and placebo arms, respectively.

**Table 5.**
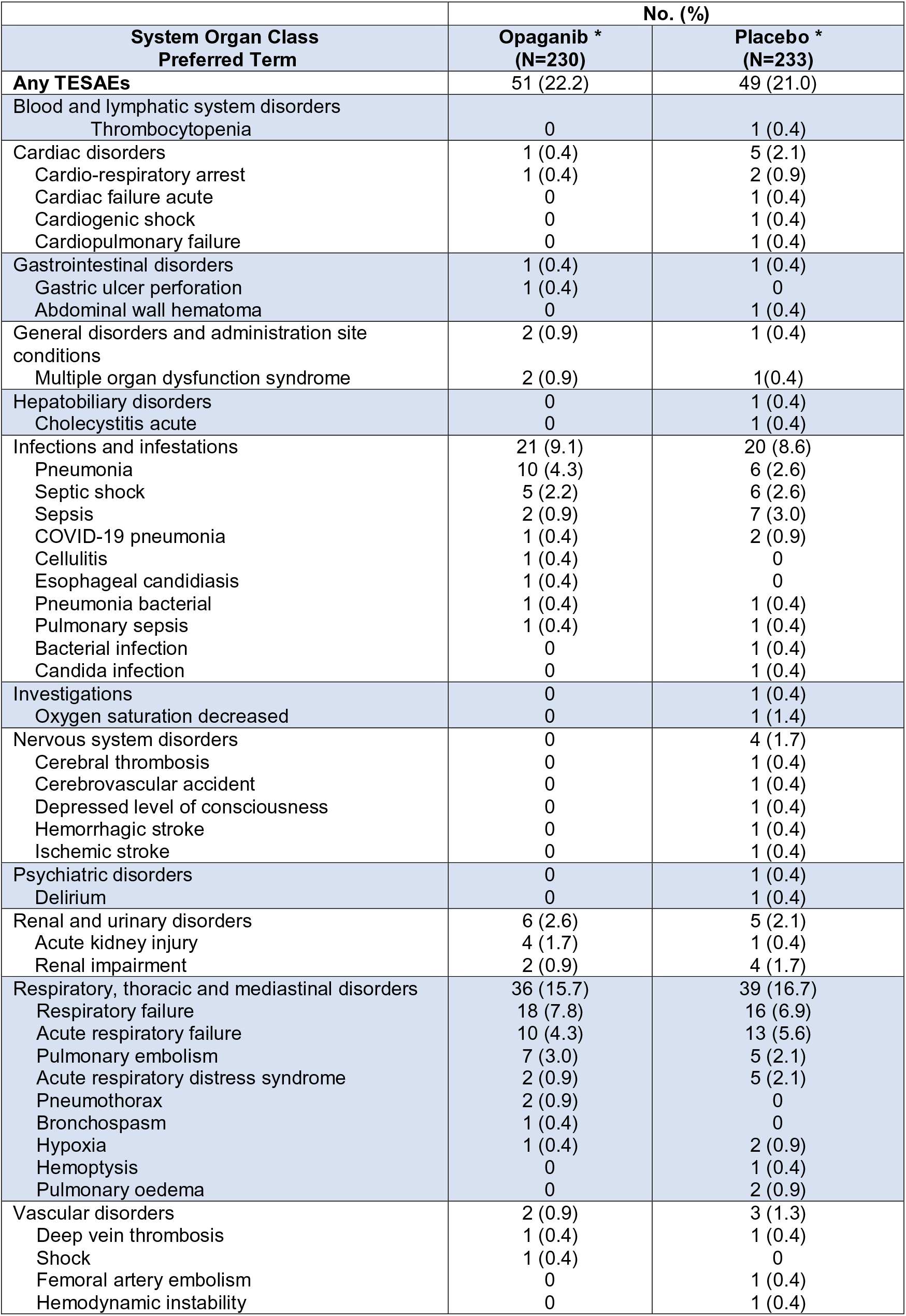
TESAE by SoC and PT by treatment group. *Patients are counted only once in each system organ class category, and only once in each preferred term category.

Thirty-nine, or 17% of all adverse events experienced in the opaganib arm were considered related to treatment. In the placebo arm, 29 (12.4%) adverse events were consider related to treatment. Psychiatric adverse events included insomnia which occurred at a frequency of greater than 5% (7.4% versus 3.9% in the opaganib and placebo arm, respectively; Table 6). Insomnia is included as an AE of special interest (AESI) and discussed with other neuro-psychiatric events in sections below. The only TESAE deemed to be related to drug treatment by the investigators while still blinded was an single incident of grade 2 change in mental status that resolved within 24 hours of the medication being withdrawn.

**Table 6.** TEAEs by treatment group and by grade (safety population). *Patients are counted only once in each system organ class category, and only once in each preferred term category. †Grade 2 event that resolved within 24 hours of study drug cessation.

| EVENT | No. (%) |  |
| --- | --- | --- |
|  | Opaganib *<br>(N=230) | Placebo *<br>(N=233) |
| Any TEAEs | 155 (67.4) | 147 (63.1) |
| Serious TEAEs | 52 (22.6) | 52 (22.3) |
| Grade 1 TEAEs Mild | 115 (50.0) | 109 (46.8) |
| Grade 2 TEAEs Moderate | 58 (25.2) | 67 (28.8) |
| Grade 3 TEAEs Severe | 32 (13.9) | 29 (12.4) |
| Grade 4 TEAEs Life threatening | 17 (7.4) | 16 (6.9) |
| Grade 5 TEAEs Death | 36 (15.7) | 40 (17.2) |
| Treatment Related TEAEs | 39 (17.0) | 29 (12.4) |
| Treatment Related TESAEs | 1 (0.4)† | 0 |
| TEAEs Resulting in Dose Reduced | 2 (0.9) | 3 (1.3) |
| TEAEs Resulting in Drug Withdrawn | 26 (11.3) | 26 (11.2) |

Premature discontinuation from the study (between day 15 and day 42, inclusively) occurred in 48 (20.9%) and 51 (21.9%) patients in the opaganib and placebo arms, respectively. Overall, there were 76 treatment emergent deaths in the study (Table E10 in the online data supplement). Most of the deaths in both groups were assessed as related to COVID-19 disease and associated complications. No deaths were assessed as being related to study drug.

Due to high mortality in a blinded review, RedHill suspended enrolment at site 114 in October 2020. The Data Safety Management Board endorsed permanent closure of this site after review of unblinded data that showed very high within-site mortality rate due to the same outcome of “worsening pneumonia” for 8/15 (53.3%) patients. After database lock it was noted that of the 15 patients enrolled at site 114, 12 patients were randomized to the opaganib arm and 3 patients to the placebo arm, all of whom were in the >60% baseline FiO_2_ subpopulation, with 7/12 patients in the opaganib arm and 1/3 patients in the placebo having an SAE of worsening pneumonia with an outcome of death. This anomaly possibly introduced a confound for both the mITT and FiO_2_>60% populations, particularly for the mortality rate outcomes in the study.

## DISCUSSION

This was a phase 2/3 multi-center randomized, double-blind, parallel arm, placebo-controlled study in 475 adult patients hospitalized with severe SARS-CoV-2 (COVID-19) pneumonia evaluating the treatment benefit of opaganib therapy vs. placebo. Primary analysis was based on the proportion of patients who no longer needed supplemental oxygen by Day 14 (end of treatment) with secondary objectives looking at improvement in other clinical outcomes such as mortality.

The pre-specified analyses in the mITT showed no significant differences with treatment, with the exceptions of shorter time to viral clearance and superior outcomes for patients receiving SoC of both remdesivir and dexamethasone as well as improved time to recovery to WHO level 1 in patients treated with opaganib. A post-hoc analysis of the data, utilizing oxygen requirements at baseline to refine the categorization of COVID-19 pneumonia severity, further justified by differences in baseline inflammatory markers and lymphocyte counts, demonstrated that in a subpopulation of patients requiring FiO_2_ (≤60%) at baseline, patients receiving opaganib had better outcomes for both primary and secondary endpoints compared to patients receiving placebo. Overall, the safety events were similar between treatment arms.

Importantly, oxygen requirement as measured by baseline FiO_2_ emerged as a potential predictor of treatment benefit in severe hospitalized patients requiring supplemental oxygen regardless of oxygen delivery device, that correlated well with the SpO2:FiO2 ratio. Post-hoc analyses of this subpopulation revealed that patients receiving opaganib had nominally superior outcomes across the primary and secondary measures, including a 61.8% reduction mortality by day 42, over patients receiving placebo. This novel metric for refining categorization of severity within the WHO level 5 population is supported by analyses of biomarkers; the difference in outcomes defined by FiO_2_ requirement correlated with baseline prognostic lymphocyte counts, inflammatory markers, and D-dimer levels. These data support FiO2 as a baseline predictor of treatment benefit within this patient population.

Considering that baseline FiO_2_ was the second highest risk factor for mortality, these data suggest that opaganib may be effective in reducing the incidence of mortality in hospitalized patients with severe COVID-19 pneumonia as indicated by FiO_2_ requirements up to and including 60%.

By contrast, in the subpopulation of patients requiring FiO_2_>60% at baseline, a difference in outcomes was not observed, indicating that the overall limited effect seen in the mITT population could be attributed to this cohort of patients. The lack of treatment effect in this subpopulation may be explained by the greater severity of the underlying COVID-19 lung disease reflected by the higher inspired oxygen requirements, lower lymphocyte counts and elevated inflammatory markers at baseline, which may, in turn suggest that there may be a threshold level for disease irreversibility.^20^

Cox regression analysis demonstrated that mortality outcome differences in the FiO_2_<60% subpopulation between opaganib and placebo were independent of potential baseline characteristics confounders: in addition, the differences in severity between the low FiO_2_ (≤60%) and high FiO_2_ (>60%) subpopulations were independent of disease duration.

As illustrated by this global study, current severity classifications by the WHO Ordinal Scale for Clinical Improvement are rather general and may not be homogeneous for a diversity of settings across global sites. As a result, the type of oxygen delivery device may not be sufficient to define COVID-19 pneumonia severity. As the objective of supplemental oxygen is to deliver sufficient FiO_2_ to the patient, it stands to reason that FiO_2_ requirement at baseline may serve as a better proxy for refining how disease severity is measured.

Overall, oral administration of opaganib treatment in this large-controlled study was shown to be relatively safe and well-tolerated. The safety profile did not indicate any new safety concerns with respect to the use of opaganib in this hospitalized patient population requiring supplemental oxygen for COVID-19 pneumonia. Generally, gastrointestinal, and neuropsychiatric disorders occurred more frequently in the opaganib arm, while respiratory disorders occurred more frequently in the placebo arm. These safety results may reflect the expected opaganib adverse events for the opaganib arm, while reflecting disease progression of COVID-19 pneumonia for the placebo arm. Except for neuropsychiatric events, TEAE of special interest were similar between treatment arms. These infrequent neuropsychiatric events occurred more commonly in patients in the opaganib arm and were mostly of mild severity.

Currently, there are limited treatment options for patients with severe COVID-19 pneumonia.^21^ The emergence of new variants strains diminishes the effectiveness of both antibodies and vaccines. While the IL6 inhibitors, like tocilizumab, were positive in just one positive trial to achieve EUA approval, they were less effective for severe patients.^22–24^ Molnupiravir and Paxlovid show varying degrees of efficacy in outpatients within 5 days of symptom onset.^8,25,26^ In our study severely ill patients were randomized a median of 11 days after appearance of symptoms. Additionally, the viral clearance data demonstrate further support for opaganib’s antiviral activity. Importantly, by addressing a far more advanced disease status and a mechanism that is host-based and should be agnostic to viral variant, opaganib has the potential to fill an urgent unmet medical need for hospitalized patients with COVID-19 without effective treatment options currently available.

## CONCLUSION

This study demonstrated, in a post-hoc analysis, a potential treatment benefit in the subpopulation of hospitalized patients with severe COVID-19 pneumonia as defined by WHO criteria and requiring relatively lower supplemental oxygen requirements, as measured by FiO_2_ and supported by lower inflammatory markers and higher lymphocyte counts at baseline. The safety profile was favorable indicating a favorable overall risk-benefit for the treatment of COVID-19. These data combined with shorter time to viral clearance indicate that opaganib may be an effective new oral therapy for COVID-19. Further studies are warranted.

## Data Availability

In general, RedHill Biopharma Ltd. adopts ICMJE requirements regarding data sharing as detailed in RedHill's Data Sharing Plan.

## ETHICS DECLARATION

The Ethics Committees in all participating sites in Brazil, Mexico, Poland, Colombia, Italy, Israel and Russia have approved the conduct of the study. Following is a list of the Ethics Committees:

Santa Casa De Misericordia De Sao

Faculdade De Medicina Do ABC/Fundacao Do ABC-FMABC

USP-Hospital Das Clinicas Da Faculdade De Medicina Da Universidade De Sao Paulo-HCFMUSP

Centro Universitario De Maringa – UNICESUMAR

Hospital Regional Hans Dieter Schmidt/SES/SC (CEP)

Universidade Do Sul De Santa Catarina - UNISUL (CEP)

Hospital Ver Cruz - HVC/MG (HVC)

Hospital De Clinicas De Porto Alegre Da Universidade Federal Do Rio Grande Do Sul - HCPA UFRGS

Universidade De Passo Fundo/Vice-Reitoria De Pesquisa E Pos-Graduacao-VRPPG/UPF

Hospital Felicio Rocho/MG

Biosafety Research Committee of the American British Cowdray Medical Center IAP

Biosafety Research Committee of the American British Cowdray Medical Center IAP

Hospital La Mision - Research Ethic Committee

The independent Ethics Committee at the Warmia and Mazury Chamber of Physicians in Olsztyn

Research Ethics Committee: Centro Medico Imbanao

Universidad Pontificia Bolivariana

Comite de Etica en Investigacion en seres Humanos Ceish

Comite de Etica de la Investigacion

National Clinics/en Alianza Con/Hospital Universitario Clinica San Rafael-Research

Ethics Committee

Comitato Etico Brianza

Ospedale Luigi Sacco-Regionale Lombardia

Comitato Etico Interaziendale, Azienda Ospedaliera Nazionale

Comitato Etico Interaziendale, A.O.U. Città della Salute e della Scienza di Torino

Shaare Zedek Medical Center Helsinki Committee

Assuta Samsom Hospital Helsinki Committee

Ziv Medical Center Helsinki Committee

Wolfson Medical Center Helsinki Committee

Galilee Medical Center, Helsinki Committee

EMMS Nazareth Hospital, Helsinki Committee

Biomedical Ethics Committee of the State Budgetary Healthcare Institution of the City of Moscow N.V. Sklifosovsky Clinica

The Independent Ethics Committee at RSBHI-Clinical Hospital No 1

I.P. Pavlov Ryazan State Med University-Local Ethics Committee

Ministry of Health for the Tver Region-State Budgetary Healthcare Institution for the Tver Region-Ethics Committee

Ministry of Health of the Russian Federation-IV Razumovsky Saratov State Medical

University of the Ministry of Health of Russia

St Petersburg State Budgetary Institution Elizavetinskaya Hospital-Bioethics Ethics Committee

Ministry of Health for Altai Krai -Local Ethics Committee

St Petersburg State healthcare Institution Municipal Hospital No 15

Yaroslavl Regional Clinical Hospital of War Veterans Ethics Committee

St Petersburg State University-Bioethics Ethics Committee

St Petersburg State Budgetary Institution Elizavetinskaya Hospital Ethics Committee

Murmansk Regiona Clinical Hospital Named After PA Bayandin Ethics Committee

St Petersburg State Budgetary Institution Elizavetinskaya Hospital Ethics Committee

Local Ethics Committee of the Saint Petersburg State Budgetary Healthcare Institution

City Pokrovskaya Hospital

Federal State Budgetary Institution Central Clinical Hospital -Ethics Committee

