## Supplementary Data for "Effect of Opaganib on Supplemental Oxygen and Mortality in Patients with Severe SARS-CoV-2 Pneumonia"

**ONLINE DATA SUPPLEMENT**

Fernando Carvalho Neuenschwander, Ofra Barnett-Griness, Stefania Piconi, Yasmin Maor, Eduardo Sprinz, Nimer Assy, Oleg Khmelnitskiy, Nikita Lomakin, Boris Mikhailovich Goloshchekin, Ewelina Nahorecka, Adilson Joaquim Westheimer Calvacante, Anastasia Ivanova, Sergey Vladimirovich Zhuravel, Galina Yurevna Trufanova, Stefano Bonora, Amer Saffoury, Ami Mayo, Yury Shvarts, Giuliano Rizzardini, Rogerio Sobroza de Mello, Janaina Pilau, Alexey Klinov, Benjamin Valente-Acosta, Oleg Olegovich Burlaka, Natalia Bakhtina, Maskit Bar-Meir, Ivan Nikolaevich Shishimorov, Jose Oñate-Gutierrez, Cristian Ivan Garcia Rincon, Tatiana Ivanovna Martynenko, Ludhmila Abrahão Hajjar, Ana Carolina Nazare de Mendonca Procopio, Krzystof Simon, Walter Gabriel Chaves Santiago, Adam Fronczak, Conrado Roberto Hoffmann Filho, Osama Hussein, Vladimir Aleksandrovich Martynov, Guido Chichino, Piotr Blewaska, Jacek Wroblewski, Sergio Saul Irizar Santana, Andres Felipe Ocampo Agudelo, Adam Barczyk, Rachael L. Gerlach, Eppie Campbell, Aida Bibliowicz, Reza Fathi, Patricia Anderson, Gilead Raday, Michal Klein, Clara Fehrmann, Gina Eagle, Vered Katz Ben-Yair, Mark L. Levitt

**METODOLOGY**

**INCLUSION AND EXCLUSION CRITERIA**

All patients were 18 to 80 years of age (inclusively), tested positive for SARS-CoV-2 infection, and were receiving supplemental oxygen as high flow, positive pressure ventilation, or non-rebreather face mask at high oxygen concentrations. Patient-reported race and ethnicity categories were collected as part of the demographic characteristics. Investigators reviewed symptoms, risk factors, and other inclusion and exclusion criteria prior to enrollment.

**SECONDARY OUTCOMES**

A total of nine efficacy secondary outcomes were evaluated. Four outcome measures involved consideration of the need for oxygen supplementation including (1) two level or greater improvement in the WHO Ordinal Scale for Clinical Improvement by day 14, (2) time until recovery as defined by improvement to a score of 3 or less on the WHO Scale, (3) time until transition to low oxygen flow via nasal cannula from high oxygen flow via nasal cannula or positive pressure ventilation at baseline, (4) the proportion of patients requiring intubation and mechanical ventilation by day 42, In addition, both time to discharge from the hospital and mortality at by days 28 and 42 following the initiation of treatment were measured. An additional outcome focused on general health including measures of the proportion of patients transitioning from a fever at baseline (>38.0°C, 100.4°F), to being afebrile (<37.2°C, 99°F) by day 14. Two additional endpoints focused on infection were (1) measures of the proportion of patients with two consecutive negative swabs for SARS-CoV-2 by PCR at day 14 and (2) the time until to two consecutive negative swabs through day 14.

**EXPLORATORY OUTCOMES**

The mean change in systemic markers of inflammation (D-dimer, cardiac troponin, C-reactive protein, lactate dehydrogenase, and ferritin) as well as lymphocyte count from baseline at day 14 were to be evaluated. Time until recovery, as defined by improvement to a score of 1 or less on the WHO Ordinal Scale for Clinical Improvement, the percentage of patients no longer requiring supplemental oxygen for at least 24 hours by day 7, and the time until a 50% reduction of supplemental oxygen requirement for the subset of patients who did not receive positive pressure ventilation (non-invasive or invasive) were all evaluated in the opaganib versus placebo arm.

**STRATIFICATION FOR TREATMENT RANDOMIZATION**

Patients were randomized using 1:1 assignment ratio to receive either opaganib added to standard of care (SoC) or matching placebo added to SoC.

Stratification was to be done based on the following criteria:

1. Whether the patient met three or more high risk parameters for COVID-19 outcomes at baseline (yes or no). These parameters included: age at screening ≥ 60 years, (yes or no); male, (yes or no); HbA1c at screening ≥ 6.5 or on active treatment with insulin or oral hypoglycemics (yes or no); hypoxemia without commensurate increased work of breathing (yes or no); known underlying chronic lung disease (yes or no); known cardiovascular disease or hypertension (yes or no); body mass index (BMI) ≥ 28.0 kg/m2 (yes or no); known renal disease (yes or no).
2. Whether standard of care treatment had established efficacy (for further details, please see Efficacious Standard of Care section below).

**STATISTICAL ANALYSIS**

Eligible patients (n=464) were randomized to double-blind treatment. An opaganib intent to treat (ITT) (n=232) and placebo standard of care (n=232) was met (figure 1 in supplemental material 3). This sample size calculation was based on powering the study to the primary analysis of patients no longer requiring supplemental oxygen for at least 24 hours by day 14. This calculation assumed an opaganib treatment success corresponding to an elimination of supplemental oxygen in at least 15% more of the patients than in the placebo arm. The 464 patients provided 90% power to detect the assumed difference in success rate, using a chi square test with a two-sided α=0.05 level of significance. This sample size calculation considered a planned non-binding futility analysis performed after ≥135 patients studied had been evaluated for the primary endpoint. Baseline characteristics are presented for the ITT population as shown in table 1. Efficacy endpoints were analyzed for the modified ITT (mITT) population (patients who received at least one dose of study drug). Safety evaluations were performed for the safety population (patients who took at least on dose of study drug). A post-hoc analysis was performed for evaluating COVID-19 severe pneumonia patients entering the study with ≤60% FiO_2_. The primary endpoint analysis included a Cochran Mantel-Haenzel test to compare the proportion of success between the two groups, using the study stratification factors as used for randomization, and corresponding stratified risk difference estimate along with 95% confidence interval.

**Efficacious Standard of Care**

A SoC treatment with established efficacy was defined by Emergency Use Authorization or full approval granted by either the US Food and Drug Administration (FDA), the European Medicines Agency (EMA) or the Medicines and Healthcare Products Regulatory Agency (MHRA). The proven effective therapies for the purpose of this study were adjusted as new data emerged and were documented and shared with study personnel regularly. The following treatments were included in the list of proven effective therapies: dexamethasone, remdesivir, COVID-19 convalescent plasma and baracitinib in combination with remdesivir.

**RESULTS**

| **Unmet Eligibility Criteria** | **n (%) ^*^** |
| --- | --- |
| Abnormal Liver Function Tests | 53 (46.9) |
| Prohibited con-meds | 45 (38.9) |
| Negative COVID-19 nasopharyngeal swab | 37 (32.7) |
| Screening QTC above eligibility criteria | 31 (27.4) |
| Intubation at baseline | 30 (26.6) |
| Table E1. Reasons for Screen Failures. ^*^Patients may have more than one exclusion criteria to fail eligibility. | |

|  | **No. (%)** | |  |
| --- | --- | --- | --- |
| **Parameter** | **Opaganib** | **Placebo** | **Outcome** |
| **Patients with an Improvement of 2 or More on the WHO Ordinal Scale Compared to Baseline by Day 14, n (%)^*^** | 145 (63.04) | 138 (59.23) |  |
| Difference (Opaganib% - Placebo%) |  |  | 3.81 |
| Percentage change (Opaganib%/placebo%*100) |  |  | +6.4 |
| 95% CI | 56.81, 69.28 | 52.92, 65.54 | -5.06, 12.69 |
| **Time to a score of ≤ 3 on the WHO Ordinal Scale, days** |  |  |  |
| Number of events, n (%) | 145 (63.0) | 138 (59.2) |  |
| Stratified Log-Rank Test, p-value ^†^ |  |  | 0.231 |
| Kaplan-Meier Median (days) ^‡^ | 11.00 | 12.00 |  |
| Estimate and 95% CI | 9.00 - 13.00 | 10.00 - 14.00 |  |
| **Time to low oxygen flow via nasal cannula (from high flow nasal cannula or CPAP/BiPAP), days** |  |  |  |
| Number of events, n (%) | 169 (73.5) | 167 (71.7) |  |
| Stratified Log-Rank Test, p-value |  |  | 0.472 |
| Kaplan-Meier Median (days), Estimate 95% CI | 5.50 (5.00 - 6.00) | 6.00 (5.00 - 7.00) |  |
| **Time to Discharge by day 14** |  |  |  |
| Number of Events n (%) | 128 (55.7) | 132 (56.7) |  |
| Stratified Log-Rank Test, p-value |  |  | 0.521 |
| Kaplan-Meier Median (days) | 13.50 | 14.00 |  |
| 95% CI | 12.00 - 15.00 | NA – NA |  |
| **Patient requiring intubation and mechanical ventilation by day 42^d^** | 53 (23.04) | 57 (24.46) |  |
| Difference (Opaganib% - Placebo%) |  |  | -1.42 |
| Percentage change (Opaganib%/placebo%*100-1) |  |  | -5.8 |
| 95% CI | 17.60, 28.49 | 18.94, 29.98 | -9.17, 6.33 |
| Intubation Without death, n (%) | 10 (4.35) | 10 (4.29) |  |
| Intubation with Death, n (%) | 27 (11.74) | 30 (12.88) |  |
| Death without intubation, n (%) | 9 (3.91) | 8 (3.43) |  |
| Early termination/missing data (alive without intubation), n (%) | 7 (3.04) | 9 (3.86) |  |
| Difference in the proportion of "Failure" between groups ^§^ |  |  | 0.701 |
| Stratified proportion difference ^ll^ |  |  | -1.51 |
| 95% CI |  |  | -9.19, 6.18 |
| Difference in Rates (Opaganib - Placebo) |  |  | -2.36 |
| 95% CI | 11.72, 21.32 | 13.86, 23.91 | -9.31, 4.59 |
| Difference in the proportion of "Failure" between groups ** |  |  | 0.488 |
| **Mortality due to any cause at day 42 (“Failure”), n (%)** | 44 (19.13) | 47 (20.17) |  |
| Difference (Opaganib% - Placebo%) |  |  | -1.04 |
| Percentage change (Opaganib%/placebo%*100-1) |  |  | -5.2% |
| 95% CI | 14.05, 24.21 | 15.02, 25.32 | -8.28, 6.20 |
| Difference in the proportion of "Failure" between groups |  |  | 0.755 |
| Table E2. Secondary outcomes for mITT population. | | | |
| **^*^**Success definition: patient who reached improvement of at least 2 points on WHO scale by Day 14 and maintained this by end of study (EOS).  ^†^p-value from Cochran Mantel-Haenzel test using the study stratification factors used for randomization, and corresponding stratified proportion difference with 95% CI.  ^‡^Estimated using the Kaplan-Meier estimator  ^§^Failure definition: any requirement of intubation and mechanical ventilation, or death without intubation, by day 42. Early termination of study (or failure to complete EOS visit) is also considered as failure.  ^ll^A Cochran Mantel-Haenszel test to compare the proportion of failure between the two groups including stratified proportion difference and its 95% CI, using the study stratification factors used for randomization  ^**^Mortality (one of the reasons that define "failure") is assessed up to and including Day 42. Any early termination/ missing survival status at EOS visit is also regarded as “failure” for the primary analysis of this endpoint | | | |

| **Parameter** | **Opaganib (N=218)** | **Placebo (N=219)** |
| --- | --- | --- |
| The time to two consecutive negative swabs for SARS-CoV-2 by PCR, at least 24 hours apart, up to 14 days |  |  |
| Number of Events | 93 (42.7) | 79 (36.1) |
| Number of Censored observations | 125 (57.3) | 140 (63.9) |
| Reasons for censoring |  |  |
| No post-baseline results available | 13 | 8 |
| Less than two results and discharged by day 5 | 7 | 7 |
| Less than two results and not discharged by day 5 | 21 | 24 |
| At least two results which are not two sequential negatives | 84 | 101 |
| Log-Rank Test Statistic^*^ | 12.58 |  |
| p-value^a^ | 0.043 |  |
| Hazard Ratio (HR) and 95% CI^†^ | 1.34 (0.99 - 1.82) |  |
| Kaplan-Meier Median Estimate and 95% CI^‡^ | 10.00 (8.00 - NA) | NA (10.00 - NA) |
| Cumulative Incidence {%}^‡^ |  |  |
| Day 7 | 42.09 | 33.25 |
| 95% CI | 35.19,49.75 | 26.89,40.65 |
| Day 14 | 54.57 | 47.69 |
| 95% CI | 46.87, 62.63 | 39.70, 56.41 |
| Table E3. The time to two consecutive negative swabs for SARS-CoV-2 by PCR, at least 24 hours apart, up to 14 days. | | |
| ^*^Analysis statistics were estimated using Log Rank test stratified by study stratification factors used for randomization. A negative (positive) statistic is associated with longer (shorter) time to the event.  ^†^Estimates and confidence intervals are obtained from Cox proportional hazards regression model with treatment group as explanatory variable and stratification factors to determine the strata levels.  ^‡^Estimated using the Kaplan-Meier estimator. | | |

|  | **No. (%)** | |  |
| --- | --- | --- | --- |
| **Parameter** | **Opaganib** | **Placebo** | **Outcome** |
| Mortality due to any cause at day 28^*^ | 2 (4.65) | 10 (21.28) |  |
| Difference (Opaganib% - Placebo%) |  |  | -16.63 |
| Percentage change (Opaganib%/placebo%*100) |  |  | -81.1 |
| 95% CI | 0.00, 10.95 | 9.58, 32.98 | -29.91, -3.34 |
| P-value |  |  | 0.024 |
| Mortality due to any cause at day 42 | 3 (6.98) | 11 (23.40) |  |
| Percentage change (Opaganib%/placebo%*100) |  |  | -16.43 |
| 95% CI | 0.00, 14.59 | 11.30, 35.51 | -30.73, -2.1 |
| P-value |  |  | 0.034 |
| Table E4. Mortality due to any cause at days 28 and 42 after remdesivir and corticosteroids SoC with or without (placebo) opaganib for mITT population.; *Mortality ("failure") is assessed by treatment day 28 or 42 (including). Any early termination/ missing survival status at EOS visit is also regarded as failure for the primary analysis of this endpoint. | | | |

| **Parameter** | **Opaganib (N=230)** | **Placebo (N=233)** |
| --- | --- | --- |
| Time to recovery as defined by improvement to a score of 1 or less on the WHO Ordinal Scale for Clinical Improvement, up to 14 days | | |
| Number of events (%) | 86 (37.4) | 65 (27.9) |
| Number of censored observations (%) | 144 (62.6) | 168 (72.1) |
| Log-Rank test statistic ^*^ | 14.89 |  |
| p-value^a^ | 0.013 |  |
| Hazard ratio (HR) and 95% CI ^†^ | 1.49 (1.08 - 2.05) |  |
| Kaplan-Meier median estimate and 95% CI | NA (NA - NA) | NA (NA - NA) |
| Cumulative incidence, % ^‡^ |  |  |
| Day 7 | 12.61 | 7.30 |
| Day 14 | 37.39 | 27.90 |

Table E5. Time to recovery as defined by improvement to a score of 1 or less on the WHO ordinal Scale for clinical Improvement in the mITT population.

CI=confidence interval; HR=hazard ratio; NA=not achieved.

^*^Analysis statistics were estimated using Log Rank test stratified by study stratification factors used for randomization. A negative (positive) statistic is associated with longer (shorter) time to the event.

^b^Estimates and confidence intervals were obtained from Cox proportional hazards regression model with treatment group as explanatory variable and stratification factors as covariates.

^‡^Estimated using the Kaplan-Meier estimator.

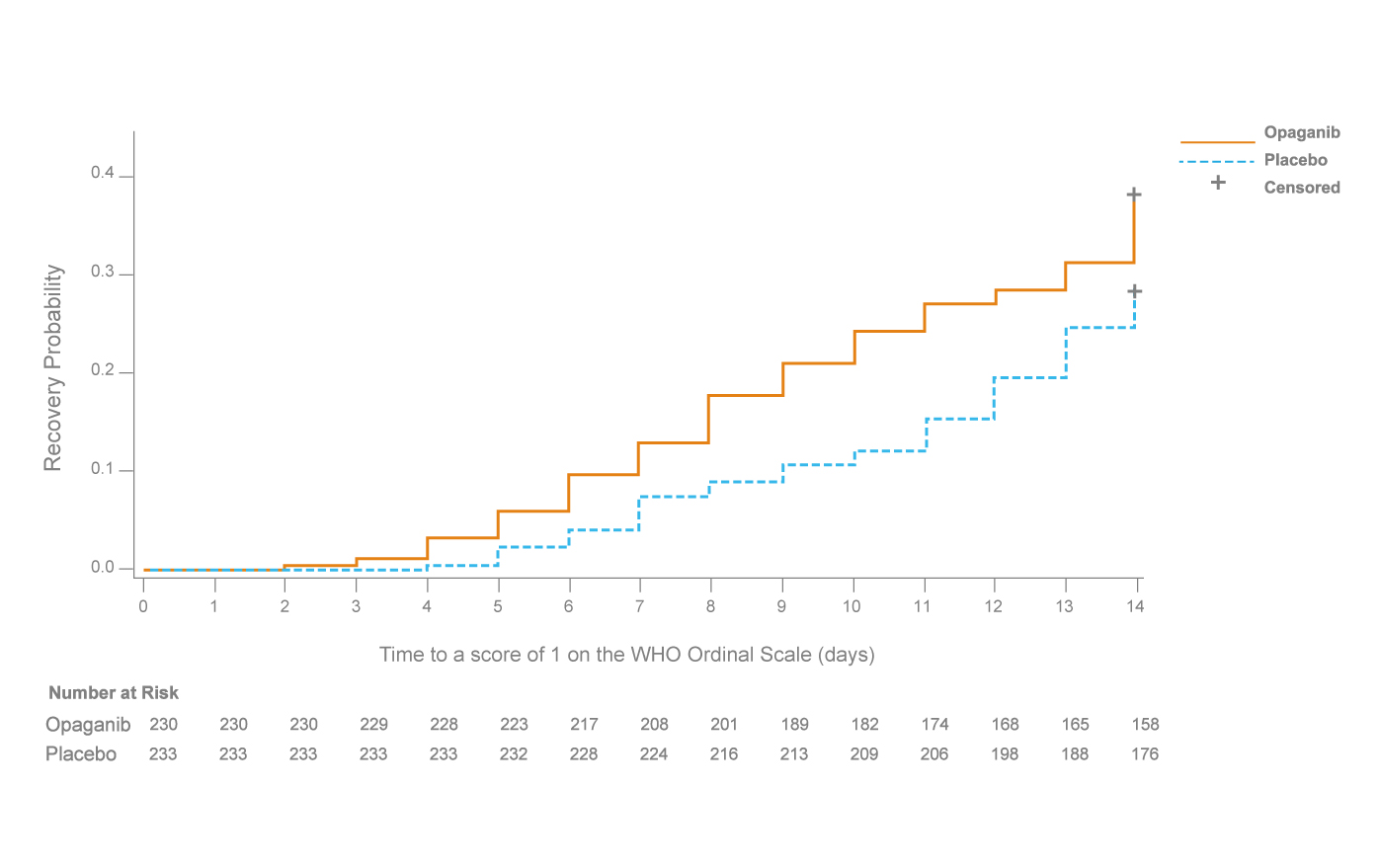

Figure E1. Kaplan-Meier curve of cumulative incidence for time to recovery as defined by improvement to as score of 1 or less on the WHO Ordinal Scale of Clinical Improvement (mITT). Subjects who were lost to follow-up or withdrew consent or died before Day 14 were censored to Day 14. Remaining subjects without the event were censored to day 14 or end of study day if occurred earlier.

| **BLHF Treated Population** **^*^** | **Statistics** | **Opaganib (N=216)** | **Placebo (N=219)** | **Outcome** |
| --- | --- | --- | --- | --- |
| **Patients No Longer Receiving Supplemental Oxygen**  **(“Success”)** **^†^** | n (%) | 126 (58.33) | 125 (57.08) |  |
| Difference in Rates (Opaganib - Placebo) | % |  |  | 1.26 |
|  | 95% CI | 51.76, 64.91 | 50.52, 63.63 | -8.03, 10.54 |
|  | p-value [2] |  |  | 0.755 |
| Stratified Success Proportion Difference (CI) ^‡^ | % (CI) |  |  | 1.45 (-7.64, 10.55) |
| “Failure” ^§^ | n (%) | 90 (41.67) | 94 (42.92) |  |
| Due to need of supplemental oxygen at Day 14 | n (%) | 70 (32.41) | 72 (32.88) |  |
| Due to death up to day 14 | n (%) | 18 (8.33) | 19 (8.68) |  |
| Due to Lost to follow up by Day 14 | n (%) | 2 (0.93) | 2 (0.91) |  |
| Due to missing status at Day 42 with prior success | n (%) | 0 | 1 (0.46) |  |

Table E6. Sensitivity analysis of the primary endpoint: percentage of patients no longer requiring supplemental oxygen, for at least 24 hours by study day 14.

N (and n) =number; CI = Confidence Interval.

^*^ ”Baseline high flow” patients defined as receiving supplemental oxygenation at baseline either by non-rebreather (reservoir) face mask, high flow nasal cannulas or non-invasive positive pressure ventilation.

^†^Success indicating that a patient no longer received supplemental oxygen for at least 24 hours by Day 14.

^‡^p-value from Cochran Mantel-Haenzel test using the study stratification factors used for randomization, and corresponding stratified proportion difference with 95%CI.

All reported p-values are nominal, two sided.

Patients can only be on one failure category.

^§^Patients who died within 42-days or were LTFU or in need of oxygen up to 42 days have been regarded as Failure.

|  | | | | | | | |
| --- | --- | --- | --- | --- | --- | --- | --- |
|  | | | **Mortality at 42 days rate (%) (Kaplan-Meier analysis)** | | | | |
| **Variable** | **# subjects with bl. data** | **cut-point for Low high groups (median)** | **High marker group** | **Low marker group** | **High-Low difference** | **CI** | **two-sided p** |
| Age | 463 | 57.000 | 26.2% | 7.0% | 19.2% | [ 12.5% , 25.8% ] | <.0001 |
| Oxygen in Gas Mix at Baseline (%) | 463 | 60.000 | 27.5% | 8.5% | 18.9% | [ 11.6% , 26.2% ] | <.0001 |
| # of risk factors | 463 | 3.000 | 25.5% | 8.7% | 16.8% | [ 9.9% , 23.7% ] | <.0001 |
| Lactate Dehydrogenase (IU/L) | 431 | 405.000 | 24.5% | 8.6% | 16.0% | [ 9.0% , 22.9% ] | <.0001 |
| Lymphocytes (10^9/L) | 457 | 0.900 | 9.1% | 23.7% | -14.6% | [ -21.4% , -7.8% ] | <.0001 |
| D-Dimer (ug/ml) | 436 | 0.578 | 21.5% | 11.0% | 10.6% | [ 3.6% , 17.5% ] | 0.0029 |
| C Reactive Protein (mg/L) | 445 | 82.800 | 21.9% | 11.5% | 10.4% | [ 3.4% , 17.4% ] | 0.0036 |
| Ferritin (ug/L) | 411 | 758.000 | 21.5% | 11.6% | 9.9% | [ 2.6% , 17.1% ] | 0.0075 |
| Oxygen Saturation at Baseline (%) | 463 | 94.000 | 11.8% | 20.1% | -8.3% | [ -15.0% , -1.6% ] | 0.0152 |
| Pulse Rate at Baseline (beats/min) | 463 | 79.000 | 19.6% | 13.7% | 6.0% | [ -0.9% , 12.9% ] | 0.0903 |
| BMI (CRF) at Baseline (kg/m^2) | 463 | 29.988 | 18.9% | 14.1% | 4.8% | [ -2.1% , 11.7% ] | 0.1754 |
| Systolic Blood Pres at Baseline (mmHg) | 463 | 129.000 | 18.9% | 14.2% | 4.7% | [ -2.1% , 11.6% ] | 0.1770 |
| Weight at Baseline (kg) | 463 | 87.100 | 17.3% | 15.3% | 2.0% | [ -4.9% , 8.9% ] | 0.5693 |
| Oxygen Flow at Baseline (L/min) | 463 | 15.000 | 17.9% | 16.1% | 1.8% | [ -5.5% , 9.0% ] | 0.6274 |
| Time from onset of symptoms to randomization (Days) | 455 | 11.000 | 17.0% | 15.8% | 1.3% | [ -5.7% , 8.3% ] | 0.7200 |
| Temperature at Baseline (C) | 463 | 36.700 | 17.3% | 16.1% | 1.2% | [ -5.8% , 8.1% ] | 0.7409 |
| Table E7. Baseline risk factors for mortality (mITT population). | | | | | | | |

**Table E7. Baseline risk factors for mortality (mITT population).**

|  | **Opaganib** | | | **Placebo** | | | |
| --- | --- | --- | --- | --- | --- | --- | --- |
| **Biomarker** | **N** | **Median** | **Q1, Q3** | **N** | **Median** | **Q1,Q3** | **p-value** **^*^** |
| Lymphocytes (10^9/L) | 116 | 0.910 | 0.69, 1.36 | 133 | 1.010 | 0.72, 1.41 | 0.2534 |
| C Reactive Protein (mg/L) | 113 | 67.100 | 26.77, 173.00 | 127 | 45.000 | 18.60, 145.60 | 0.1910 |
| Ferritin (ug/L) | 104 | 727.600 | 381.65, 1383.45 | 124 | 592.300 | 368.02, 1201.20 | 0.3025 |
| D-Dimer (ug/ml) | 111 | 0.499 | 0.17, 1.17 | 129 | 0.380 | 0.18, 1.04 | 0.6619 |
| Lactate Dehydrogenase (IU/L) | 106 | 359.400 | 300.00, 507.00 | 124 | 368.750 | 287.55, 536.34 | 0.5522 |
| Table E8. Biomarker distribution (medians, 1st and 3rd quartile) by treatment arm (FiO2≤60% subpopulation). | | | | | | | |
| * Comparing opaganib vs. placebo arms for imbalances. Baseline troponin was collected in 20% of the patients and therefore is not presented. | | | | | | | |

|  | | | | | |
| --- | --- | --- | --- | --- | --- |
|  | **Adjusted Mortality at 42 days Analysis** | | | | |
|  | **Mortality rates** | | **Opaganib-Placebo difference** | | |
| **Potential confounder** | **Opaganib** | **Placebo** | **Difference** | **lower 95% CI for difference** | **upper 95% CI for difference** |
| Study Site Identifier | 4.6% | 10.9% | -6.3% | -12.5% | -0.1% |
| # of risk factors :<=/> overall median | 4.7% | 11.2% | -6.4% | -12.9% | -0.0% |
| # of risk factors | 4.6% | 11.2% | -6.6% | -12.9% | -0.3% |
| Age :<=/> overall median | 4.6% | 11.5% | -6.9% | -13.4% | -0.4% |
| Age | 4.5% | 11.7% | -7.2% | -13.7% | -0.6% |
| Pulse Rate at Baseline (beats/min) :<=/> overall median | 4.5% | 11.7% | -7.2% | -13.7% | -0.7% |
| Cardiovascular Disease (Curr) | 4.4% | 11.7% | -7.3% | -13.7% | -0.8% |
| Male (Curr) | 4.5% | 11.9% | -7.4% | -14.2% | -0.7% |
| Positive for COVID at Screening | 4.4% | 11.9% | -7.5% | -14.1% | -0.8% |
| Time from onset of symptoms to randomization (Days) :<=/> overall median | 4.5% | 12.1% | -7.7% | -14.4% | -0.9% |
| Renal Disease (Curr) | 4.3% | 12.1% | -7.8% | -14.4% | -1.1% |
| Smoking Status | 4.7% | 12.4% | -7.8% | -14.8% | -0.7% |
| Oxygen Saturation at Baseline (%) :<=/> overall median | 4.4% | 12.1% | -7.8% | -14.5% | -1.1% |
| Weight at Baseline (kg) :<=/> overall median | 4.4% | 12.2% | -7.8% | -14.5% | -1.0% |
| Three or More Risk Factors (Curr) | 4.3% | 12.1% | -7.8% | -14.3% | -1.3% |
| Country | 4.3% | 12.1% | -7.8% | -14.5% | -1.2% |
| Oxygen in Gas Mix at Baseline (%) :<=/> overall median | 4.3% | 12.2% | -7.8% | -14.6% | -1.1% |
| Chronic Lung Disease (Curr) | 4.3% | 12.2% | -7.9% | -14.6% | -1.1% |
| HbA1c (Curr) | 4.3% | 12.2% | -7.9% | -14.6% | -1.2% |
| Temperature at Baseline (C) :<=/> overall median | 4.4% | 12.3% | -7.9% | -14.7% | -1.1% |
| Systolic Blood Pres at Baseline (mmHg) :<=/> overall median | 4.3% | 12.2% | -7.9% | -14.5% | -1.3% |
| Fever at Baseline Flag | 4.3% | 12.2% | -7.9% | -14.6% | -1.2% |
| Hypoxemia (Curr) | 4.3% | 12.3% | -8.0% | -14.7% | -1.3% |
| BMI Greater or Equal 28 | 4.2% | 12.4% | -8.2% | -14.9% | -1.5% |
| D-Dimer (ug/ml) :<=/> overall median | 3.6% | 11.8% | -8.2% | -14.8% | -1.6% |
| Lactate Dehydrogenase (IU/L) :<=/> overall median | 3.9% | 12.2% | -8.3% | -15.0% | -1.6% |
| BMI (CRF) at Baseline (kg/m^2) :<=/> overall median | 4.3% | 12.6% | -8.3% | -15.2% | -1.4% |
| Effective SOC ^*^ | 4.5% | 13.1% | -8.5% | -15.7% | -1.4% |
| Oxygen Flow at Baseline (L/min) :<=/> overall median | 4.2% | 12.8% | -8.6% | -15.5% | -1.7% |
| C Reactive Protein (mg/L) :<=/> overall median | 4.4% | 13.1% | -8.7% | -15.7% | -1.7% |
| Lymphocytes (10^9/L) :<=/> overall median | 4.0% | 13.0% | -9.0% | -15.7% | -2.3% |
| Ferritin (ug/L) :<=/> overall median | 2.9% | 13.2% | -10.3% | -17.1% | -3.5% |
| Table E9. Mortality sensitivity analysis for handling early study discontinuation 42 days for mITT with baseline FiO_2_≤60% adjusted for potential baseline confounder.  Adjusted mortality and their difference are calculated based on the method of Direct Adjusted Survival Curves' as performed by SAS PHREG procedure.  In this analysis, the confounder is entered into the model as explanatory variable and treatment group as strata variable.  Each potential confounder is analyzed separately.  Age and number of risk factors are handled both as a continuous variable and as binary variable </>median.  Overall medians are based on the overall mITT population.  ^*^ Effective Standard of Care was predefined during the study and include: glucocorticoids, remdesivir and hyperimmune plasma for COVID-19. | | | | | |

|  | **No. (%)** | | | |
| --- | --- | --- | --- | --- |
| **System Organ Class   Preferred Term** | **Opaganib mITT * (N=230)** | **Opaganib with site 114 removed * (N=218)** | **Placebo**  **mITT * (N=233)** | **Placebo with site 114 removed * (N=230)** |
| Any TEAEs | 36 (15.7) | 29 (13.3) | 40 (17.2) | 39 (16.9) |
| Cardiac disorders  Cardiac failure acute  Cardio-respiratory arrest  Cardiogenic shock  Cardiopulmonary failure | 0  0  0  0  0 |  | 4 (1.7)  1 (0.4)  1 (0.4)  1 (0.4)  1 (0.4) |  |
| General disorders and administration site conditions  Multiple organ dysfunction syndrome | 2 (0.9)  2 (0.9) |  | 1 (0.4)  1 (0.4) |  |
| Infections and infestations  Pneumonia  Septic shock  Pulmonary sepsis  Sepsis  COVID-19 pneumonia  Pneumonia bacterial | 16 (7.0)  10 (4.3)  4 (1.7)  1 (0.4)  1 (0.4)  0  0 | 9 (4.1)  3 (5.2) | 12 (5.2)  3 (1.3)  3 (1.3)  0  4 (1.7)  1 (0.4)  1 (0.4) | 11 (4.8)  2 (0.8) |
| Investigations  Oxygen saturation decreased | 0  0 |  | 1 (0.4)  1 (0.4) |  |
| Nervous system disorders  Cerebrovascular accident  Haemorrhagic stroke  Ischemic stroke | 0  0  0  0 |  | 3 (1.3)  1 (0.4)  1 (0.4)  1 (0.4) |  |
| Respiratory, thoracic, and mediastinal disorders  Respiratory failure  Pulmonary embolism  Acute respiratory distress syndrome  Acute respiratory failure  Pneumothorax  Pulmonary oedema | 18 (7.8)  9 (3.9)  3 (1.3)  2 (0.9)  2 (0.9)  2 (0.9)  0 |  | 18 (7.7)  8 (3.4)  2 (0.9)  4 (1.7)  3 (1.3)  0  1 (0.4) |  |
| Vascular disorders  Femoral artery embolism | 0  0 |  | 1 (0.4)  1 (0.4) |  |
| Table E10. TEAEs with an outcome of death, SoC and preferred term by treatment group with and without site 114. *****Patients are counted only once in each system organ class category, and only once in each preferred term category. Worsening of COVID-19 pneumonia was coded to COVID-19 pneumonia. | | | | |
